# Non-household environments make a major contribution to dengue transmission: Implications for vector control

**DOI:** 10.1101/2024.01.08.24301016

**Authors:** Víctor Hugo Peña-García, A. Desiree LaBeaud, Bryson A. Ndenga, Francis M. Mutuku, Donal Bisanzio, Jason R. Andrews, Erin A. Mordecai

**Affiliations:** Department of Biology, Stanford University, Stanford, CA, USA; School of Medicine, Stanford University, Stanford, CA, USA; Kenya Medical Research Institute, Kisumu, Kenya; Department of Environmental and Health Sciences, Technical University of Mombasa, Mombasa, Kenya; RTI International, Washington, DC, USA

**Keywords:** Dengue, agent-based model, non-household environments, vector control

## Abstract

The incidence of Aedes-borne pathogens has been increasing despite vector control efforts. Control strategies typically target households, where Aedes mosquitoes breed in household containers and bite indoors. However, our study in Kenyan cities Kisumu and Ukunda (2019-2022) reveals high Aedes abundance in public spaces, prompting the question: how important are non-household (NH) environments for dengue transmission and control? Using field data and human activity patterns, we developed an agent-based model simulating transmission across household (HH) and five NH environments, which was then used to evaluate preventive (before an epidemic) and reactive (after an epidemic commences) vector control scenarios. Our findings estimate over half of infections occur in NH settings, particularly workplaces, markets, and recreational sites. Control efforts in NH areas proved more effective than HH, contradicting the current global focus. Greater reductions in dengue cases occurred with early, high-coverage interventions, especially in NH locations. Additionally, local ecological factors, such as uneven water container distribution, influence control outcomes. This study underscores the importance of vector control in both household and non-household environments in endemic settings. It highlights a specific approach to inform evidence-based decision making to target limited vector control resources for optimal control

## 1 Introduction

Vector-borne diseases are a group of infections caused by pathogens such as parasites, bacteria, and viruses that are transmitted by biting arthropods; together, they put 80% of world’s population at risk [1]. Dengue virus, transmitted by *Aedes* mosquitoes, is among the most important vector-borne diseases because of the close relationship with human environments and its large and growing burden [2, 3]. Dengue is estimated to cause around 400 million infections globally per year [4, 5] and recent trends show an increase in cases annually [6].

This increase in dengue transmission is occurring despite implementation of control activities in endemic settings. While there is some uncertainty about the effectiveness of actual control strategies due to lack of reliable evidence [7], some authors argue that vector control measures are inadequately implemented [8–10], and others add that an integrated, community-focused control requiring multisectoral, multi-disciplinary engagement, and community participation sustained in time is necessary [11]. As a result, one of the main issues to be addressed is the improvement of vector control programs [12].

The design of vector control strategies should be as effective as possible while minimizing the required costs, time, and human resources. In search of this efficiency, vector control strategies have predominantly focused on households (HH) [13]. The rationale behind this assumption is that people spend more time in household environments than in any other structure, while sharing the same space with cohabitants and biting, breeding vectors [14]. As a result, most vector control guidelines exclude non-household (NH) locations as targets of interventions [14–16]. Yet, recent evidence suggests a larger role of NH environments in infection risk [17–21], which was supported by a significant reduction of cases reported during the COVID-19 pandemic lockdowns, when people spent more time in households and less time outside of them [22, 23].

To better understand the role of NH environments in dengue transmission, we developed an agent-based model of dengue transmission and calibrated it to data on mosquito breeding places, abundance, and human activity space in various environments from two Kenyan cities [24]. With the model, we estimated the relative contribution of both HH and NH environments to dengue transmission and evaluated the outcome when control strategies are focused in either or both types of environments under two possible scenarios: preventive and reactive control.

## 2 Methods

### 2.1 Model overview

#### 2.1.1 Simulated populations and environments

The aims of this work were 1) to identify the contribution of HH and NH to the total number of dengue infections and 2) to evaluate the outcome of different vector control strategies on dengue transmission in urban areas. To do so, we developed an agent-based model incorporating data and conditions from Kenyan cities of Kisumu (located at the western part of the country next to Lake Victoria) and Ukunda (coastal city at the eastern part of the country). DENV circulation and endemicity has been described in these cities for a long time [25], where higher levels of dengue has been reported for Ukunda than Kisumu [26], spanning a wide range of endemicity levels within these populations.

To develop the model, we created two synthetic populations representing each of the Kenyan cities. To do this, we considered the number of inhabitants per household for each city according to information provided by reports of 2019 Kenya Population and Housing Census [27], which is also in accordance with previously reported household occupancy information for both cities [24], where the mean number of inhabitants per household is 4.6 for Kisumu and 7.3 for Ukunda. For tractability and scalability, we set the size of synthetic populations to be around 20,000 individuals (final size was 20,172 for Ukunda and 20,160 for Kisumu), so the number of households was set accordingly to both size of human population and the mean number of house inhabitants.

Following the aim of this work, we additionally created a synthetic NH environment. For this, we considered information related to the presence of water containers in these locations and available data about the presence of people at different urban spaces (See Supplementary material and Data Obtaining section) to define five different types of NH environments: workplaces, schools, religious spaces (representing churches, mosques, etc.), markets (including shopping places of any kind), and recreational spaces (grouping any place where people attend for entertainment or gatherings like bars, nightclubs, parks, etc.). The number of workplaces and schools were defined according to information extracted from previous reports informing the average number of either workers [28] (rounded to 19) or students [29] (rounded to 360) for each of the respective environments. Unfortunately, for the remaining NH structures, there was no available information related to their proportion or density within cities. By considering survey results from local populations, we defined their density as one market and recreational place for every 30 houses and one religious space for every 50 houses.

We used data on proportions of water-holding containers in HH and NH environments that we previously published [24]. With those proportions, each structure was assigned a specific number of water containers, each with an assigned size based on our data on the size distribution of containers [24].

Each individual of the synthetic population was assigned an age following the proportions reported in the Census of 2019 [27]. Each individual was also assigned an occupation being either student, worker, both, or none (for example, toddlers or retired) following the general student and worker age reported in Kenyan Quarterly Labour Force Report (2021)[28] and Basic Education Statistical Booklet (2019) [29]. We assumed an initial baseline prevalence of dengue of 0.08% estimated from previous studies reporting age-structured seroprevalence [30] with an incidence rate per year estimated as 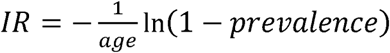.

#### 2.1.2 Population dynamics

The model was configured to simulate on a daily basis what happens in each structure where humans and vectors are present and hence, infections can take place. The presence of humans at a given location depends on the type of structure, where households, workplaces and schools are daily-attending locations, and religious, markets, and recreational places are daily-randomly assigned attendings. In this sense, for each structure falling into the first category, the same individuals attend daily. For the latter three types of structures, the number and selection of individuals occurs randomly on a daily basis (See Supplementary material). Finally, we also included movement among HH with a probability of 0.1 for a given HH to receive a non-resident individual each day. To consider mosquito movement, we turned to information from studies that release large numbers of mosquitoes, which may promote mosquito movement, which have shown that roughly 90% of mosquitoes are recaptured either within 30 meters of the release point or even in the same house [31–33]. For this reason, we assumed that mosquito movement between buildings for an already established subpopulation is negligible (no movement). Instead, in the model the human movement is the primary driver of virus spread.

When a given structure has mosquitoes and humans there is a chance of transmission if any of them is infected. The probability of a human being bitten in a given structure depends on the number of both mosquitoes and humans and the time that humans spend in the structure. For simplicity, each structure was assigned with a specific number of hours for people to spend that is defined according to the type of structure. Data to define the number of hours per structure was estimated based on fieldwork conducted in the same study cities (See Data Acquisition section and Supplementary material).

Mosquito population dynamics are not determined at a city-wise but at a structure-wise level, recognizing that different patterns of transmission are obtained when space fragmentation is considered [34]. To do this, we considered that the main limiting driver of mosquito population growth occurs at the larval stages [35, 36] and hence depending on the breeding place availability, their volume, and larval density. Accordingly, we developed a density-dependent function to estimate the number of individuals in the next discrete time (day) describing the larval survival as a function of the density of mosquito immature stages and temperature, as follows:

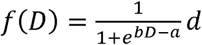

Where

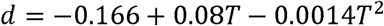

In this equation, *a* and *b* are calibrated values (see Supplementary material), *d* is a term that relates temperature (*T*) with the remaining terms in the equation and *D* is the larval density expressed as the ratio of the number of larvae and the number of liters of water available for breeding in the structure. The number of larvae and mosquito mortality rates are also temperature-dependent following the functions described previously [37–39] (see Supplementary material for details related to implementation of functions in this work).

#### 2.1.3 Infection dynamics

For a susceptible mosquito that bites an infected human, it can be moved from susceptible to exposed stage according to temperature-dependent vector competence and later be moved to infectious stage with temperature-dependent EIP (extrinsic incubation period) following functions previously described (Table S2) [37]. Infectious mosquitoes remain in this stage until death, for which the rate also depends on temperature using a previously published function (Table S2) [37].

Humans that are bitten by infected mosquitoes are moved to latent stage where they remain for five days. At the end of this period, they are moved to the infectious stage lasting seven days before moving to a recovered stage. Since the model does not explicitly simulate the circulation of DENV serotypes, we considered a period of complete protection before returning to a susceptible state to account for multiple serotype infections. Based on Sabin’s classic works of experimental infections [40, 41]), complete heterotypic protection can be lost after 3 months after exposure so we set a complete heterotypic immunity lasting for 100 days.

The type of structure where each infection takes place and the date were recorded. Each computational run considers a temporal window of 731 days comprising between January 1^st^ of 2020 until December 31^st^ of 2021. Data are temporarily grouped yielding the number of cases happening every week. Results are expressed as median and interquartile range (IQR) of infections among 400 runs. The model was coded in Julia language v1.8 and all simulations were run on Sherlock cluster at Stanford University (Stanford Research Computing Center).

### 2.2 Data acquisition

Information related to the number of containers in different environments was obtained from fieldwork performed between 2020 and 2022 in both study sites and previously reported in detail [24]. Briefly, 400 m^2^ urban areas were sampled with four different strategies targeting different stages of mosquitoes: ovitraps to obtain information on eggs and egg-laying females, container surveys to obtain information of larval stages and container availability, Prokopack aspirators to obtain information related to adults, and BG-Sentinel to obtain information from breeding place-searching females.

Information on sample location was recorded including the type of environment (HH or NH), number of water containers and their size category, the *Aedes* positivity status, and number of house inhabitants was also recorded when applicable.

Serological data was obtained only for purposes of calibrating the model (Supplementary Material). Data consisted of sero-incidence estimates based on individuals at all ages yielding a negative serology followed by a positive test during a follow-up examination six months later. Though the study contains data from 2014 to 2022, only those individuals recruited during the same temporal window of this study was considered for calibration, i.e. from 2019 to 2022. Enrollment of individuals [42, 43] and serological methods [26] has been previously described.

Temperature data was collected in both cities by using temperature data loggers (HOBO®), and the daily average was calculated during the entire simulated period comprising from January 1^st^ of 2020 until December 31^st^ of 2022.

### 2.3 Human movement survey

We carried out a semi-structured interview (SSI) to gather information about people’s movement routine in Kenyan settings from November 1^st^ to December 2^nd^, 2021. SSIs have been previously employed to gather data on routine human movement in other settings [44]. The survey was carried out in two community cohorts corresponding to both Kenyan cities included in this work, i.e., Kisumu and Ukunda. Both cohorts are part of an ongoing longitudinal study [26]. We interviewed 201 individuals in Ukunda and 243 in Kisumu, carefully selected to represent the gender and age group distribution of their respective cohorts. The SSI included questions aimed at capturing commonly visited locations during weekly activities. Participants were asked to list locations, besides their households, such as workplaces, markets and shops, schools, and religious places that they usually visit in a week. Participants also provided an estimated amount of time spent in each location per week. The SSI was conducted by trained local technicians who provided an overview of the study. Questionnaires targeting children unable to respond by their own were completed by their guardians. For subjects under 18 years of age capable of answering the questionnaire, we obtained permission from their guardians. All locations listed by participants were georeferenced by the survey team, either by collecting GPS ground points or by gathering coordinates through Google Maps. The questionnaire used in this study is available as Supplementary material.

### 2.4 Vector Control Strategies Assessed

The strategies tested in this model are focused on the reduction of vector populations. The intensity of the control was quantified as the reduction percentage of water-holding containers available as mosquito breeding places. Container elimination was evaluated under two control scenarios termed “preventive” and “reactive”. The preventive control scenario refers to the elimination of containers on day 0 preceding the start of an epidemic. To ensure a proper start of the epidemic at the designated time, there were no infections happening in the two weeks preceding it. On the other hand, the reactive control scenario refers to the elimination of containers after the rise in cases defining the start of an epidemic. Because reactive control initiatives can take some days to be implemented for several reasons (planning, resource allocation, recruitment, among other stages), we considered 1, 50, and 100 days after the start of the epidemic as different reaction times (the day when control was accomplished, additional results for control implemented after 250 days are included in Supplementary material).

For both control scenarios, the effort of the control strategies was quantified as percentage of water containers removed. Given that our goal was also to quantify the contribution of urban HH and NH environments in the total number of cases, we additionally evaluated the effectiveness of control when it is focused only on one or both of these categories of environments. In addition, we also included the outcome of the control when it is focused on large (those with a volume higher that ten liters) or small (with a volume less than ten liters) containers to provide insights on different mosquito population-related parameters and their effects on transmission and subsequent control effectiveness.

## 3 Results

### 3.1 Infections in non-household environments are higher than expected

During a period of 731 days (i.e. two years), the model yielded a median of 784 (Interquartile Range [IQR]: 350 – 1,557) infections in Kisumu and a median of 3,971 (IQR: 2,680 – 5,445) in Ukunda (figure 1). By explicitly quantifying the number of infections happening in different urban environments, the model estimated a slightly higher proportion of infections taking place in NH structures, accounting for around 57.4% (IQR: 47.7 – 58.2) of infections from Kisumu and 56.3% (IQR: 53.1 – 57.4) of infections from Ukunda. Workplaces were the most common NH site of infections, accounting for 77.1% (IQR: 54.7-99) in Kisumu and 30.9% (IQR: 28.8 – 32.4) in Ukunda. Markets and shopping locations accounted for 9.3% (IQR: 0 – 18.8) and 27.4% (IQR: 26.2 – 29.2) of NH infections for Kisumu and Ukunda, respectively. Recreational locations accounted for 10.9% (IQR: 0.0 – 19.4) of NH infections in Kisumu and 21.4% (IQR: 20.3 – 21.8) in Ukunda. Finally, religious places and schools had the lowest number of infections, accounting for 2.7% (IQR: 0.0 – 7.1) and 0% (no infections recorded) for Kisumu and 18.9% (IQR: 17.9 – 19.1) and 1.3% (IQR: 0.0 – 5.1) for Ukunda, respectively.

**Figure 1:**
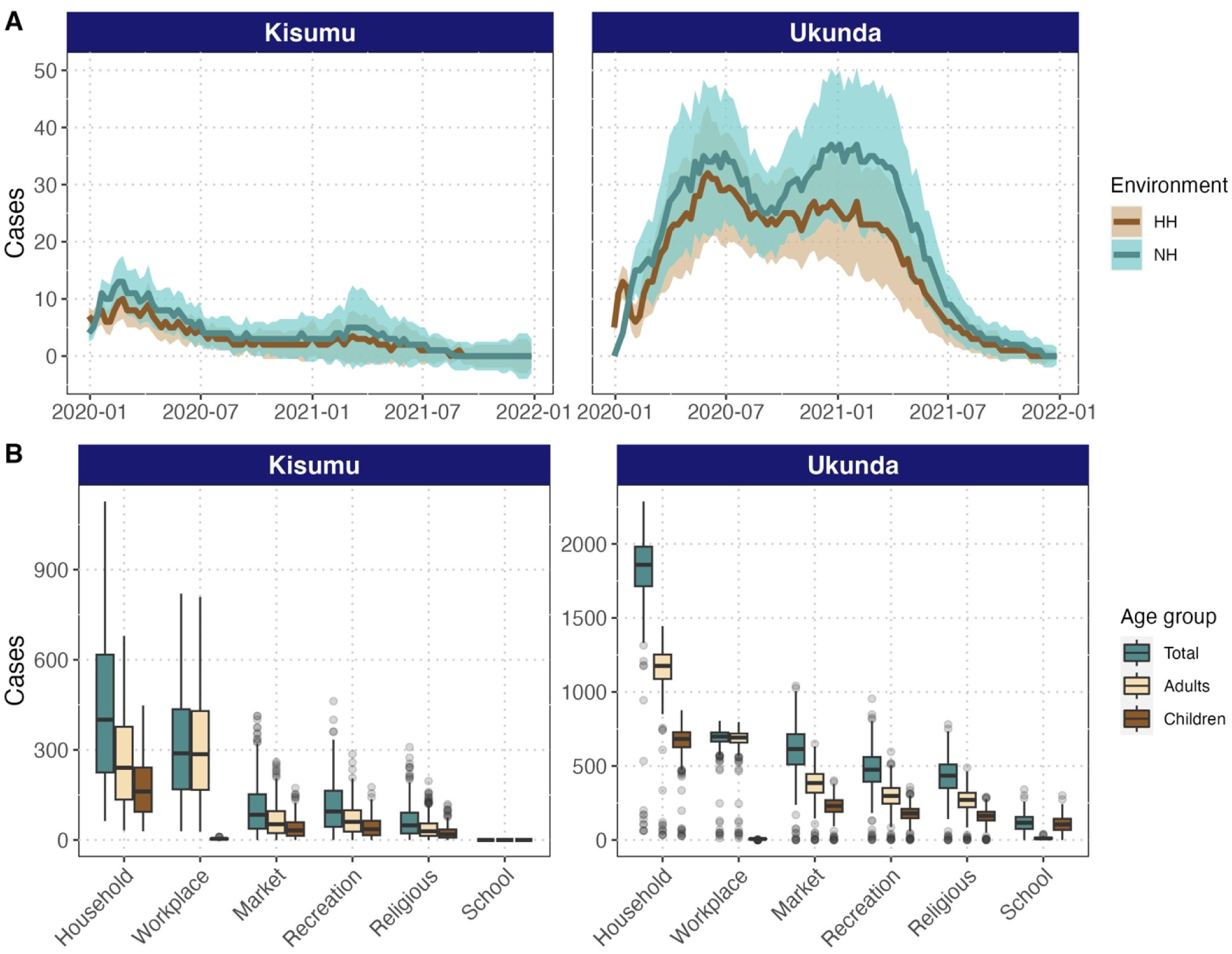
HH and NH environments contribute nearly equally to dengue transmission. Number of infections (y-axis) over time (x-axis) by environment, age, and city. The number of infections recorded along the simulated two-year period by environments and cities is depicted in panel A (shaded areas represents inter-quartile range, IQR). Panel B shows the distribution of infections by including both the total number and by age group (considering children those individuals 15 years old or younger and adults those older than 15) for the six environments considered for each city.

In addition, we wanted to understand how infection risk for children (15 years old and younger) and adults (those older than 15 years) was distributed in different environments. Though infections in schools only took place in Ukunda, in that setting a higher proportion of infections in children is happening in schools, as expected, while a higher proportion of infections in adults happen in workplaces. Following schools, the model predicted the highest proportion of infections in children happening in households followed by recreational places for both cities (figure 1).

### 3.2 Differential effectiveness in preventive control among urban environments

NH environments provide a powerful lever for vector control, especially when vector control was preventative. Control in NH was more effective than control in HH: for example, considering 50% control intensity in Kisumu, HH-only control reduced cases by 54.2% (IQR: 10.1 - 73.3) while NH-only control reduced cases by 68% (IQR: 48.4 - 80.4), and controlling both reduced cases by 76.6% (IQR: 61.3 - 85.7). In Ukunda, the difference is even more dramatic: 35.6% (IQR: 2.2 - 66.2) reduction in cases from control in HH containers alone, 84.6% (IQR: 62.9 - 95.4) reduction for NH containers alone, and 93.2% (IQR: 80.4 - 97.5) reduction for control in both HH and NH (figure 2).

**Figure 2:**
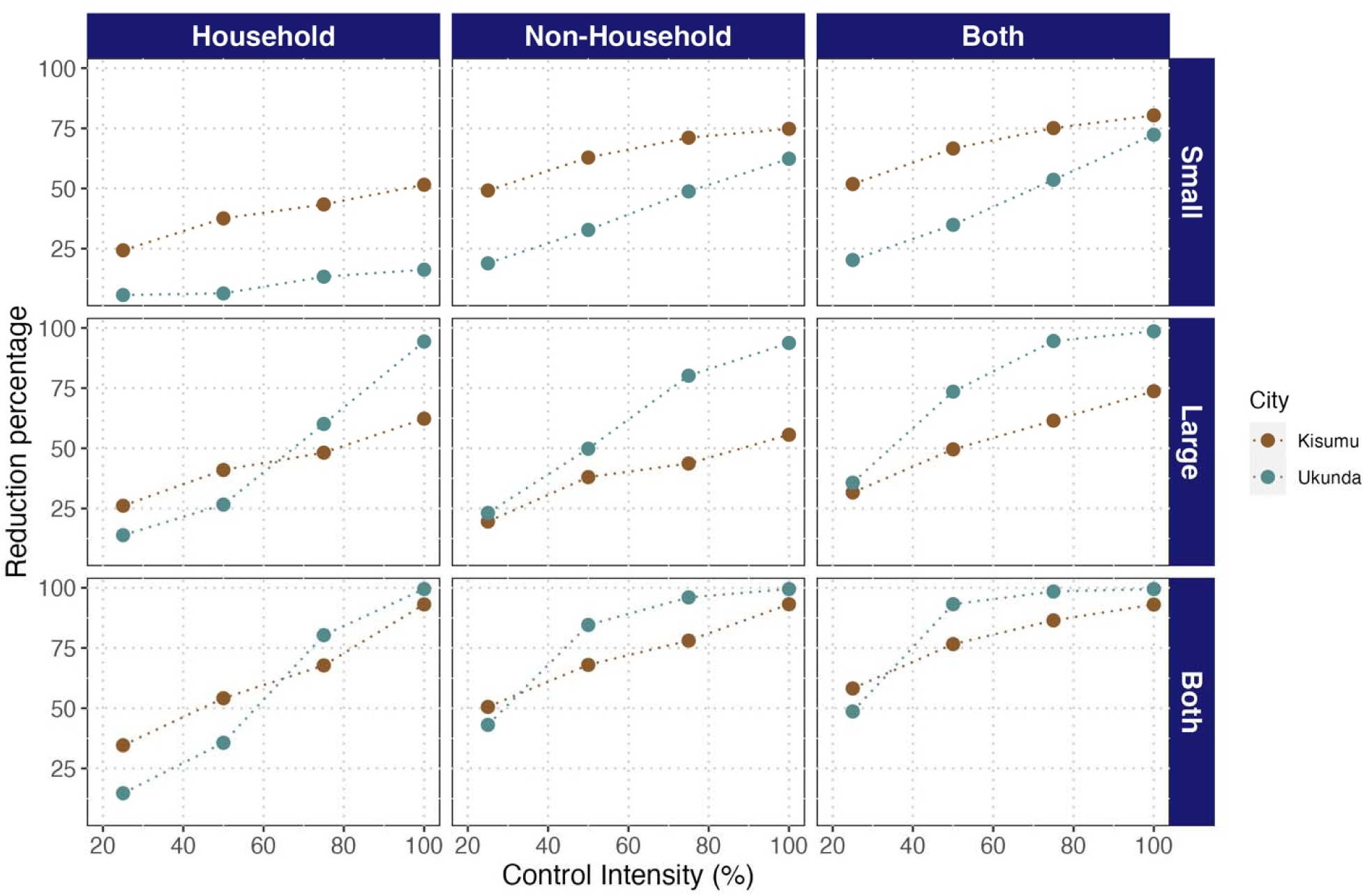
NH environments are equally or more effective than HH environments for dengue control across cities, container sizes, and control intensities, which combine to determine the most effective strategy. Effectiveness of vector control strategies evaluated under the preventive scenario. Effectiveness is expressed as reduction in percentage of dengue cases compared to the epidemic size with no vector control interventions. The vector control strategies vary according to control intensity (number of containers eliminated) and the target environment (Households, Non-Households or both) and container size (small for water containers with capacity less than ten liters, large for containers with capacity greater than ten liters, or irrespective of size).

On the other hand, the importance of container size differed between cities, where small containers (less than ten liters volume) were more important for Kisumu while removal of large containers (more than ten liters volume) yielded a greater reduction of cases in Ukunda. Thus, when 50% control intensity is applied on both environments, a reduction of 66.6% (IQR: 45.6 - 79.6) of cases can be seen when only small containers are removed while a 49.5% (IQR: 0.0 - 70.3) reduction resulted from removing only large containers in Kisumu. Nevertheless, a greater reduction is observed when the control is done irrespective of the size of the container with a reduction of 76.6% (IQR: 61.3 - 85.7) in the same city. On the other hand, in Ukunda, removing only small containers led to a 34.9% (IQR: 0.6 - 66.1) of cases reduction versus a 73.5% (IQR: 43.7 - 94.8) reduction when large containers are targeted. Similar to Kisumu, the greatest reduction in cases resulted when control is performed on all container types, with a 93.2% (IQR: 80.4 - 97.5) case reduction in Ukunda (figure 2). It is worth noting that the number of removed containers differs among cities when targeting different container sizes (figure S5). Intriguingly, the number of removed containers is consistently higher for different control intensity at HH environments (figure S5) than NH. In this sense, targeting NH should render less effort than HH and hence more efficiency.

### 3.3 The effectiveness difference among HH and NH is consistent between preventive and reactive strategies

As expected, the greatest reduction in cases is observed when vector control strategies are implemented sooner to the beginning of the epidemic showing timing to be as important as the environment. For example, considering a 50% control intensity, the effectiveness declines from 74.3% (IQR: 60.7 - 85.8) when control is implemented at day one to 44.3% (IQR: 20.5 - 62.7) when control is implemented at day 100 for Kisumu, and from 89.1% (at day one, IQR: 75.3 - 94.9) to 63.6% (at day 100, IQR: 44.9- 78.8) in Ukunda, representing an increase in effectiveness of 30% and 25.5% for Kisumu and Ukunda, respectively. By increasing the intensity to 100%, the effectiveness shifts from 90.2% (IQR: 86.3 - 96.1) at day one to 52.9% (36.1 - 66.8) at day 100 in Kisumu, and from 98.5% (IQR: 97.8 - 99.0) at day one to 80.4% (IQR: 72.5 - 82.9) at day 100 in Ukunda. On another hand, when we examined the change in the effectiveness by increasing the control intensity at day 50, it goes from 68.8% (IQR: 57.5 - 77.4) to 44.99% (IQR: 8.3 - 65.0) when intensity is shifted from 100% to 25% in Kisumu. In Ukunda, the change is sharper, going from 91.8% (IQR: 88.6 - 94.4) effectiveness at 100% of control intensity to 37.7% (IQR: 7.25 - 62.9) effectiveness at 25% of control intensity (figure 3).

**Figure 3:**
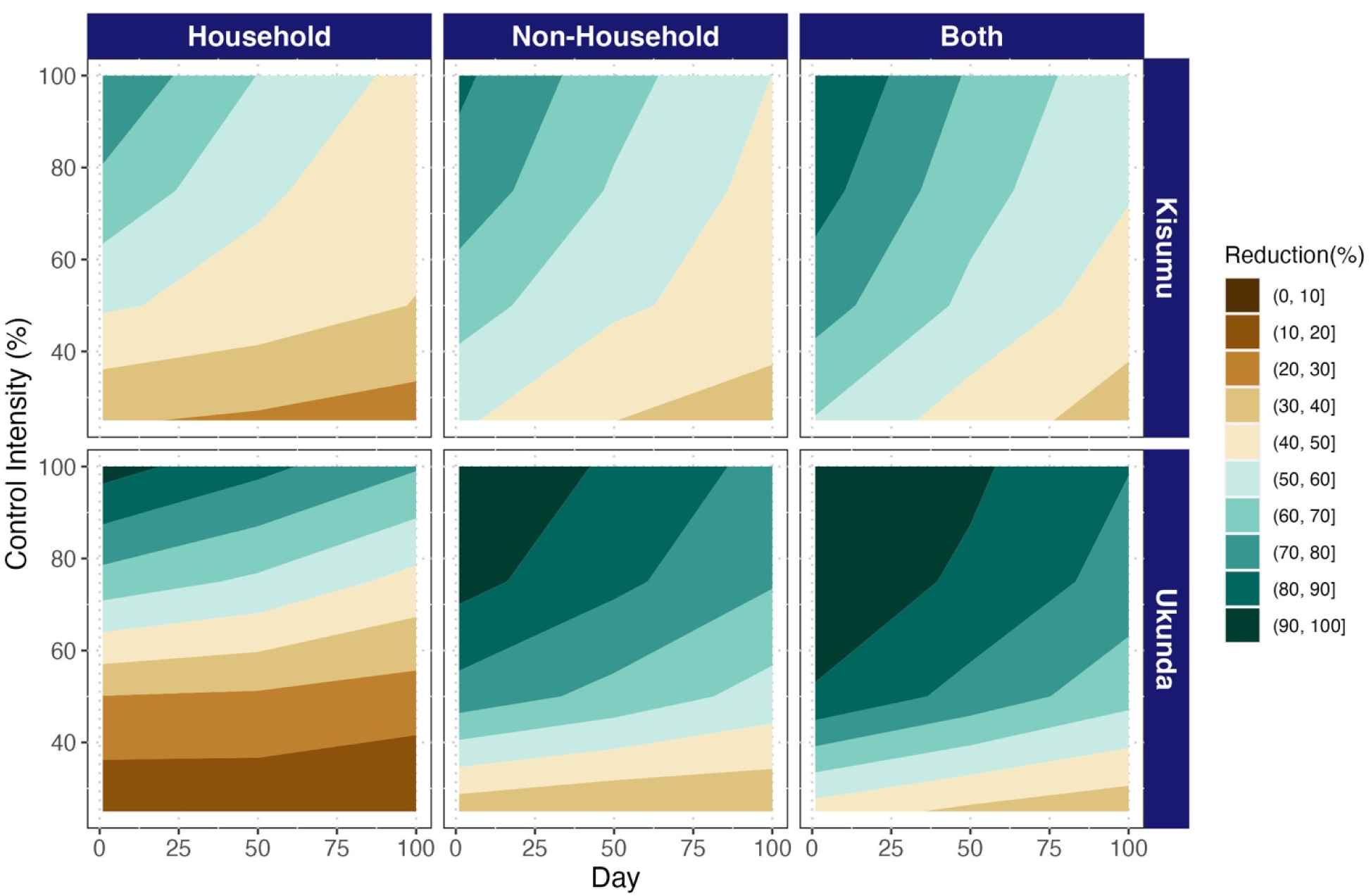
Reactive control is more effective in non-household or all environments combined than household environments alone, regardless of timing, intensity, and city. Effectiveness is expressed as the reduction in the percentage of dengue cases compared to the epidemic curve with no vector control interventions. The vector control strategies vary according to control intensity (number of containers eliminated; y-axis), the target environment (Households, Non-Households or both; panel columns); the day of implementation after the beginning of the epidemic (x-axis); and city (panel rows).

Like in the preventive scenario, for reactive control the highest reduction is achieved when control is applied in HH and NH environments, capturing their unique contributions to transmission. However, when control is applied in only one or the other, control targeted to NH environments was more effective than that targeted to HH (figure 3 and figures S2-S3).

## 4 Discussion

Recent field work reported a high number of vectors in NH environments than HH in the Kenyan cities of Kisumu and Ukunda, suggesting potentially high risk for *Aedes*-borne viruses transmission in these environments [24]. Consequently, this work expands our knowledge about the total burden of transmission varying based on human activity space within HH and NH environments as well as others like container type and density between cities. Here, we developed an agent-based model that incorporates field data on vector occurrence and abundance, container density and type, and human activity space across age structure in Kisumu and Ukunda to explore the consequences of NH environments for dengue transmission. Specifically, we tested the hypothesis that dengue vector control could be improved by extending it to NH spaces. The model supports this hypothesis, demonstrating that the contribution of NH spaces to transmission is as high or higher than HH (figure 1), and though the higher efficiency is achieved by focusing only in NH environments (figure S5), the higher reduction of cases was reached by controlling vectors in both NH and HH environments (figures. 2-3).

By looking beyond the number of infections in NH, our model suggests that these environments act as spreaders of the virus among households. In this way, while the number of new infections per household are limited by household size, the high levels of movement of individuals in NH environments provides a source of new infections and transmission spreading among households. In line with this, some previous modeling work had suggested the importance of movement of people in dengue transmission [45–48]. It is very likely that this role is also mediated by intermediate spaces among households like those defined in this work as NH environments, such as workplaces, schools, social spaces, religious spaces, and marketplaces, where the presence of vectors had already suggested a role in transmission [17, 18, 21].

Among the five categories of NH spaces, workplaces contributed most to transmission (figure 1). This is not the first work suggesting such a large contribution. Previous work developed on a Zika outbreak in Singapore by Prem and colleagues predicted an even higher proportion of infections, with an estimated of 64% (at least 51%) of infections happening at workplaces [49]. Besides HH and schools, workplaces are the locations where individuals spend most of their time, which increases the probability of being bitten by a mosquito (see Supplementary material for details of parameterization). For these spaces, we considered an average of 19 people per workplace (Supplementary Material), which is higher than the average number of inhabitants per household and hence increases the probability of having an infected individual at a given timepoint. By contrast, in schools, which have a considerably higher average number of students of 360 (higher than household and workplaces, see Supplementary material), while the probability of having an infected person at any given time is high, the probability of a mosquito biting the infected individual among the entire student population is low, and the density of mosquitoes in schools is not high enough to compensate for this low per-person biting probability. This is especially true in Kisumu, where low overall incidence explains the lack of infections in schools, while the high incidence in Ukunda increases the presence of infected individuals in schools and hence the probability of human-to-vector infections and subsequent spread. Considering the population size with which we worked (see Methods section), previous reports of infection risk in schools [18, 19], and the vulnerability of children that congregate in schools, our data suggests that this type of environment still represents some level of infection risk and its inclusion in vector control activities is necessary. This is particularly true as this is the main NH environment where children are at risk for dengue exposure.

In line with this, the proportion of infections in children and adults in different environments likely reflects the age structure of individuals visiting these locations. In this model, we assume that the epidemic starts in a fully susceptible population; accounting for age-structured variation in pre-existing immunity would possibly alter the risk scenario for children [50]. This assumption is realistic for a new serotype invading the population at least 100 days after the most recent epidemic.

The scientific literature supports the presence of significant risk in spaces other than HH in other parts of the globe [18–21]. Some of these previously identified spaces that were not evaluated in this work include abandoned and open spaces and hotels, because we did not have data available on time spent in these locations. Accordingly, it is possible that a slight increase of NH infections is still to be quantified by considering those environments.

Results under the preventive and reactive scenarios yielded similar results: higher effectiveness is achieved when control considers only NH compared to only HH, and a combined approach that includes HH and NH is most effective. This points to an important disconnect between our results and current vector control practices, which primarily focus on HH spaces and neglect NH vector control [14–16]. Scientific literature related to vector control household-focused interventions is extensive [13, 51], while there are no interventions designed to be focused on both. Though we cannot definitively attribute the lack of successful dengue control to transmission in NH settings, our data-driven model suggests that this could be an important part of the problem, and we advocate for future studies in other locations studying this phenomenon as well as potential approaches to NH vector control.

Our model includes a novel mosquito density-dependent function that allows us to realistically model vector population dynamics from a larval perspective and at the scale of individual containers (as described by McCormack and colleagues [34]), which also allows us to estimate the relative importance of different container size in transmission and control (see Supplementary material). By using the function, our results suggest that the relative importance of different types of containers is city-specific since it depends on the frequency of these containers across cities. Accordingly, small containers in Kisumu are much more frequent than large containers (See Supplementary material) so slightly higher effectiveness is achieved when control is targeting only them. The situation is different in Ukunda, where a higher effectiveness is observed when focusing on both types of containers, where control targeting large containers is more effective as this type of container is more productive [52, 53]. These results suggest that the effectiveness of container-focused vector control depends on a trade-off between the productivity of containers and their frequency, where large containers are more productive but less frequent than small containers. Though the model is using fieldwork-derived data, the purpose of this model is understanding the relative contribution of different urban environments. We think a separate analysis should be done in order to specifically evaluate the relative importance of different container types and the best approach to take advantage of their differential distribution to achieve the best outcome.

This model is meant to realistically represent the transmission conditions in both cities but has some limitations. Though agent-based models are excellent for capturing heterogeneity and variation within populations, especially when rich data are available for parameterization, our model does not estimate some other potential sources of variation like mosquito movement or variability in time in the number of breeding places. Likewise, the addition of other types of buildings beyond the six types we included in this study might provide a more comprehensive perspective of urban locations where transmission can be happening, like those described previously by our team [24]. Our estimates of movement are mainly based on the time people spend in given locations, but other potential sources of variability were not included like intra-urban distances [47, 54], decrease of mobility due to illness [55], and travel to other urban centers and rural areas. Movement data were collected using semi-structured interview (SSI, see Methods section), which relies on people’s recollection. Consequently, the collected data may be influenced by recollection bias, a common limitation of SSIs. Recollection bias could lead participants to list only highly-visited locations, potentially overlooking places that are less frequently visited. Additionally, the time spent in each location may be affected by participants’ different senses of time, which are linked to their individual characteristics.

Unfortunately, data related to density of mosquitoes in specific NH locations were not available or did not match with information collected from the human movement data survey. Some assessed spaces reported by Peña-García [24] having mosquitoes like “open spaces”, “gardens” or “banana plantation” were not reported by people as places where they spend any time. Likewise, NH categories included in the model like “workplaces” can group some of the categories reported by Peña-García. For these reasons, the initial conditions for the mosquito dynamics were the same for all NH buildings in the model. In this way, differences related to different NH locations are mainly due to the number of people attending the locations and the time spent in these. It is important to mention that mosquito positivity status per location and their number of containers were randomly assigned considering the total variability found for NH locations in the work of Peña-García [24] (Supporting Methods).

In conclusion, the results of this work suggest not only that NH locations are important in dengue transmission, but that vector control activities will be inefficient at reducing dengue burden if these spaces are not included. When exploring in detail the NH locations, differential risk depends on the number of people and the time they spend in these places, which can make some age groups particularly vulnerable to infection at given locations and certain locations critical for certain age groups. Equally, because cities differ in their abundance and distribution of container sizes, comprehensive vector control approaches that focus on multiple types of containers across HH and NH spaces are necessary to break chains of transmission. Through our model, we provide evidence-based insights for new directions aimed at designing new vector control strategies that can make limited resources be used in optimal control activities.

## Ethics

This work specifically did not collect data from human samples or surveys. However, we did acquire information from other works obtaining such information. For those, ethical approval and oversight for data collection were obtained from the Institutional Review Board of Stanford University (IRB 31488), as well as the Kenya Medical Research Institutes (KEMRI SSC 2611) and Technical University of Mombasa Ethical Review Committee (TUM/ERC EXT/004/2019).

## Data accessibility

The data, files and code are publicly available through the GitHub digital repository https://github.com/vhpenagarcia/ABM_dengue.

Supplementary material is available online.

## Declaration of AI use

We have not used AI-assisted technologies in creating this article.

## Data Availability

All data produced are available online at github.com/vhpenagarcia/ABM_dengue

## Acknowledgements

We would like to acknowledge the teams that performed fieldwork and collected the data on which this work was based on in both Kisumu and Ukunda cities: Joel Omaru Mbakaya, Samwuel Otieno Ndire, Gladys Adhiambo Agola, Paul S. Mutuku, Said L. Malumbo, and Charles M. Ng’ang’a.

## Authors’ contributions

V.H.P.G: Conceptualization, Formal Analysis, Investigation, Methodology, Software, Investigation, Validation, Writing – original draft, Visualization. A.D.L.: Conceptualization, Writing – Review and Editing, Supervision, Project administration, Funding acquisition. B.A.N.: Investigation, Data curation. F.M.M.: Investigation, Data curation. D.B.: Investigation, Data curation, Writing – review and editing. J.R.A.: Conceptualization, Methodology, Visualization, Writing – review and editing, Supervision. E.A.M.: Conceptualization, Methodology, Visualization, Writing – review and editing, Supervision.

