## Supplementary Material for "Non-household environments make a major contribution to dengue transmission: Implications for vector control"

Mombasa, Kenya

^5^ RTI International, Washington, DC, USA

*Contributed equally

### Supplementary Methods description

#### Synthetic populations

Human population was created jointly with the number of households. According to 2019 Kenyan population and housing census, Kisumu and Ukunda human populations have a size of 397,957 and 77,686, respectively; and average housing size was settled at 4.6 and 7.3 [1], respectively. For simplicity and to keep computational needs low, we created a population of about 20,000 individuals (smaller than actual populations) by defining a number of houses of 2,767 for Ukunda and 4,412 for Kisumu. The average number of house inhabitants was used as a lambda parameter in a Poisson distribution to define the size of each of the households for both populations.

Following the age proportions reported by the same census, we used a discrete non-parametric distribution implemented in Julia’s package Distributions to assign age to each individual. Following Quarterly Labour Force Report (2021) [2] and Basic Education Statistical Booklet (2019) [3], which reports the age for both students and workers we used such information to assign to each individual a main occupation to be student, worker or none of them. Extracting information from these reports, we set the average number of students in schools as 360 and the average number of workers per workplace as 19, so the number of schools and workplaces was defined considering the total number of students and workers in each city. Then, we assigned a school or workplace to which each individual attend to. To do this, we used a discrete uniform distribution to ensure that the number of individuals was evenly distributed among the available workplaces or schools by assigning to each of these structures a numbering code. To define the number of churches, markets (or shopping places) and recreational facilities, we determined to have 1 every 50 houses for churches or 1 every 30 houses for markets and recreational.

During fieldwork, data related to container size was classified as small (lower than 10 liters), medium (between 10 and 25 liters) and large (larger than 25 liters). However, we grouped both medium and large containers in the same category (10 liters) to limit the growth of mosquito subpopulation inside the HH or NH building. When the structure only had small containers, their water capacity was assigned by using a Poisson distribution with parameter λ=1 and lower-truncated in 1 to avoid containers with less than 1 liter of water to meet the nature of data obtained from fieldwork. The number of positive containers per building was simulated according to data extracted from fieldwork previously reported [1]. Parameters used to assign a given location a container-positivity status and the number of containers are shown in Table S1.

**Table S1: Mosquito population parameters used to establish structure-level populations in both cities.** Proportions were coupled into binomial distributions as probabilities. Two types of proportions are shown in the table: proportion of buildings having water containers and the proportions of buildings having specific size containers. Shape is the parameter used in Pareto distribution to determine the number of containers of a given size in each container-having building.

| **City** | **Env** | **Prop*** | ***Aedes* pos** | **small** | | **medium** | | **Large** | |
| --- | --- | --- | --- | --- | --- | --- | --- | --- | --- |
|  |  |  |  | **prop**** | **shape** | **prop**** | **shape** | **prop**** | **shape** |
| Kisumu | HH | 0.403 | 0.269 | 0.903 | 1.800 | 0.295 | 2.181 | 0.065 | 16.831 |
|  | NH | 0.307 | 0.406 | 0.868 | 0.844 | 0.162 | 1.206 | 0.025 | 3.034 |
| Ukunda | HH | 0.838 | 0.122 | 0.483 | 3.077 | 0.567 | 1.754 | 0.1 | 16.232 |
|  | NH | 0.626 | 0.198 | 0.6 | 1.668 | 0.242 | 3.366 | 0.2 | 3.641 |

* Proportion of buildings having water containers potentially serving as mosquito breeding places

** Proportion of container-having buildings that have containers of the specified size

#### Description of the model

##### Mosquito dynamics

Mosquito population dynamics are based on the internal conditions of each building. Population growth depends on the water availability according to density-dependent function described above (See methods section) and temperature, following model developed by Mordecai and colleagues [4].

**Table S2: Implementation of mosquito-related functions to model population dynamics traits.** Functions were extracted from previous works originally fitted by Mordecai [4] and later modified and used by Huber [5] and Caldwell [6]. Functions can be either Brière [*cT(T-T_min_)(T_max_-T)^1/2^*] or quadratic [*c(T-T_max_)(T-T_min_)*].

| **Trait** | **Estimation function** | | | | **Use** |
| --- | --- | --- | --- | --- | --- |
|  | **function** | **c** | **Tmin** | **Tmax** |  |
| Man biting rate (*a*) | Brière | 2.02e^-04^ | 13.35 | 40.08 | $N_{B}\sim Bin\left( Nm,a \right)$* |
| Mortality rate (*µ*) | Quadratic | -1.48e^-01^ | 9.16 | 37.73 | *N_D_*~𝐵𝑖𝑛(𝑁𝑚, 𝜇)^+^ |
| Probaility of infection of a mosquito (*c*) | Brière | 4.91e^-04^ | 12.22 | 37.46 | $vc=b\cdot c$^†^  𝐸~𝐵𝑖𝑛(*N*_𝑏𝑖𝑡_,𝑣𝑐)^‡^ |
| Probaility for a mosquito to become infectious (*b*) | Brière | 8.49e^-04^ | 17.05 | 35.83 |  |
| Parasite development rate (*PDR*) | Brière | 6.65e^-05^ | 10.68 | 45.90 | $inf\sim Bin\left( E,PDR \right)$^¥^ |
| Eggs per female (*EFD*) | Brière | 8.56e^-03^ | 14.58 | 34.61 | 𝐸𝑔𝑔𝑠~𝑃𝑜𝑖𝑠𝑠𝑜𝑛(𝐸𝐹𝐷) |
| Mosquito development rate (*MDR*) | Brière | 7.86e^-05^ | 11.36 | 39.17 | 𝑟=𝑀𝐷𝑅∙𝑝𝐸𝐴∙𝑓(𝐷)  𝐸𝑚~𝐵𝑖𝑛(*N_L_*,𝑟)^§^ |
| Egg-to-adult survival probability (*pEA*) | Quadratic | -5.99e^-03^ | 13.56 | 38.29 |  |

* *N_B_*, the number of mosquitoes at a given location biting that specific day.

^+^ *N_D_*, the number of deaths happening in a given day at a specific location.

^†^ Parameters *b* and *c* are the components of vector competence (*vc*).

^‡^ *E*, number of exposed mosquitoes, *N_bit_* refers to those mosquitoes that bit an infected individual, so they are moved to exposed infection status with a probability *vc*.

^¥^ *inf* refers to the moment when a mosquito is moved to infectious stage which is determined by a rate *PDR*

^§^ *Em*, number of emerged mosquitoes, *N_L_* is the number of larvae at a given location.

At first instance, we developed a density-dependent function to determine the population growth at a building-based level as opposed to city-wide level. This function determines the probability of larval survival based on the estimated density according to availability of water that can be used for mosquito breeding and the temperature-dependent number of eggs laid by female. We modified the equation from the original one provided by Walker and colleagues [7] and originally tested by using data extracted from literature [8-12]. Because the function provides an estimation of larval survival based only on density, it would provide a faster increase of population at colder temperatures since the number of eggs laid by female decrease with temperature [4]. In this way, less density would be expected and hence higher survival and number of mosquitoes. Since it is not realistic, we parameterized an additional term accounting for a quadratic term of temperature (*d* term in density-dependent function, methods section) that maximizes the probability around 29°C while decreasing steadily as temperature deviates from this value. In order to obtain a stochastic output of mosquito traits, mosquito parameters were coupled to a binomial or Poisson distribution to generate random numbers, as indicated in table S2.

To use the function jointly with functions described in table S1, we estimated the number of larvae considering that simulations were run on a daily basis and mosquito development takes several days. We estimated the number of females that fed and laid the eggs with a probability of

$$p_{fed}=1-e^{-\frac{a}{\mu}}$$

Where *a* is the biting rate and *µ* is the mortality rate. That probability was used to generate the number of females that fed in a given location by using a binomial distribution. The total number of larvae was estimated like the product of total larvae that fed and the number of eggs per female previously estimated as table 1.

##### Human mobility

We parameterized human mobility as the number of hours that an individual spends at different locations in one day. For simplicity, each location was assigned a specific number of hours for people to spend to. This assigned value depended on type of location and according to information collected from fieldwork in the study sites.

The data related to number of hours that each person spends at a given location were collected as a categorical variable comprising three classes: “more than 8 hours”, “between 4 and 8 hours” and “less than 4 hours”. To transform this information into numbers, we transformed the frequencies into a range of hours for people to spend to. To do this we took as a base the more frequent class. When the most frequent was the category “more than 8 hours” we set a maximum number of hours to 12 and when the more frequent class was “less than 4 hours”, we set a minimum number of hours to 1. The remaining limits of the range were established according to frequency of people falling in the adjacent category (figure S1). Once the range was established for all types of environment and cities, the number of hours for a specific location was assigned following a uniform distribution. The data for the number of hours for type of environment can be found on table S3.

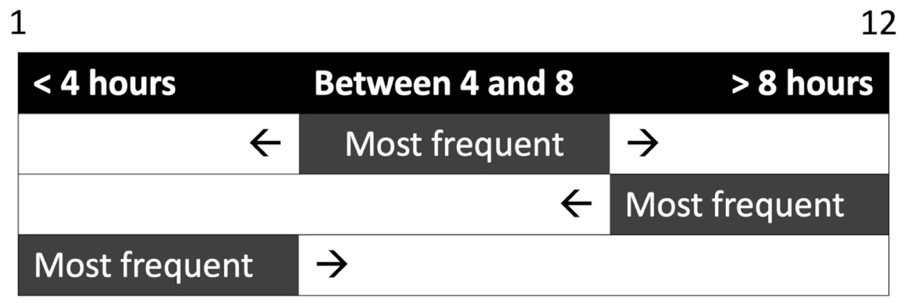

**Figure S1: Schematic representation of hours-per-day range determination for every structure according to categorical data collected from fieldwork.** The figure represents the three scenarios where any of the three categories is the most frequent. The arrows represent the direction on which the range is extended according to the frequency in which the adjacent categories are present in the survey. The minimal and maximal number of hours for the categories “less than 4 hours” and “more than 8 hours”, respectively, are also represented as 1 and 12 at the top.

**Table S3: Frequency for categories of the variable “number of hours” and the parameterized range for all environment types in both cities.** Frequencies were used to obtain a minimum and maximum number of hours for each location. Those parameters were used into a uniform distribution to define for each structure the number of hours that people spend in them every simulated day.

| **City** | **Environment** | **Hours class** | | | **Distribution parameters** | |
| --- | --- | --- | --- | --- | --- | --- |
|  |  | **Less than 4** | **Between 4 and 8** | **More than 8** | **Min** | **Max** |
| Kisumu | Household | 4 | 54 | 115 | 6.8 | 24.0 |
|  | School | 2 | 10 | 44 | 7.3 | 12.0 |
|  | Workplace | 11 | 55 | 56 | 3.6 | 9.8 |
|  | Market | 294 | 28 | 1 | 1.0 | 4.3 |
|  | Religious | 49 | 54 | 15 | 2.3 | 8.5 |
|  | Recreational | 43 | 53 | 33 | 2.7 | 9.0 |
| Ukunda | Household | 21 | 73 | 92 | 6.4 | 12.0 |
|  | School | 4 | 17 | 31 | 6.7 | 12.0 |
|  | Workplace | 4 | 51 | 32 | 3.8 | 9.5 |
|  | Market | 76 | 63 | 3 | 1.9 | 8.1 |
|  | Religious | 0 | 14 | 9 | 4.0 | 9.6 |
|  | Recreational | 6 | 4 | 5 | 2.4 | 9.3 |

In addition, we included in the model a marginal movement of people between households. To include this, we considered a probability of 0.1 for a given household to receive a non-inhabitant individual on a given day (*visitor*, from here on). When a given household is receiving a *visitor*, it is randomly selected among the population irrespective of the infection status and spends a random number of hours between 1 and 3.

Our model considers that humans attend on a daily basis household and either schools or workplaces. But the presence of individuals in places like markets (or shopping in general), recreational or religious is not following a regular pattern, so these are the spaces for interaction with other individuals whose school or workplaces are not shared. To include this randomness in the model, for each of these locations the number of individuals and “who” was randomly generated on a daily basis. To do this, every day we let the model choose the number of individuals to be one between 10 and 70, following a discrete uniform distribution implemented in the package Distributions in Julia software. Then this number of subjects were selected from among all possible subjects in the city.

##### Infection dynamics

Instead of iterating on individuals, we set the model to iterate on structures, which can have or not an associated mosquito sub-population. However, the relevant locations for the model are those in which at least one infected mosquito or human is present and hence an infectious event can happen. On these, the number of biting mosquitoes depends on the total number of mosquitoes (*Nm*) in the structure and temperature, according to function described in table S2. For those mosquitoes willing to bite, the probability of being fed (*p(b)*) depends on the probability of having an encounter with a human at any time of the day, which in turn depends on the number of hours in the location (*t_h_*) and the number of people (denoted as *Nh* in next equation) visiting such location the same day. In this way, the probability is given by

$$p\left( b \right)=1-e^{-\frac{t_{h}}{24}\times N_{h}}$$

with this probability, we were able to generate a total number of bites happening in the location by coupling it into a binomial distribution, as follows

$$b\sim Bin\left( V_{b}, p\left( b \right) \right)$$

In this equation, *b* is the number of bites and *V_b_* is the number of vectors biting Once there is a number of bites generated, three scenarios can be possible: that there are infectious humans that can infect vectors, that there are infected vectors that can infect humans, or both. On the first two scenarios, a different process is involved, and in the latter, the two processes involving the first two scenarios are happening.

When the structure has infected vectors, the number of infectious bites is determined by the number of infectious vectors following a hypergeometric distribution, as follows

$$V_{Ib}\sim Hyper\left( V_{I},V_{S},b \right)$$

Where *V_Ib_* is the number of infectious mosquitoes biting, *V_I_* are the number of infectious mosquitoes in that location the same day, *V_S_* are the number of non-infected mosquitoes and *b* is the total number of bites happening that day on that location. When there are infectious mosquitoes biting, then the model randomly chooses the individual being bitten among those having presence in the location and evaluates whether the individual is either infectious or latent. In either case, there is no new infection happening. Otherwise, if the individual is susceptible, a new infection happens, and the infection status of the individual is changed to latent.

In the case when there are infectious individuals, we generated the number of new infected mosquitoes based in in a first stage on the probability for a susceptible mosquito to having an encounter with an infected human in the same space assuming to be independent events, denoted as *p(I_h-v_)*, as follows

$$p\left( I_{h-v} \right)=\left( \frac{H_{I}}{N_{h}} \right)\left( \frac{V_{S}}{N_{v}} \right)$$

Where *H_I_* and *V_S_* are the number of infected humans and susceptible mosquitoes, respectively, and *N_h_* and *N_v_* are the total number of humans and mosquitoes, respectively, in the same location that specific day. The probability was used into a binomial distribution considering the number of bites previously estimated. Once there is a given number of human-to-mosquito infectious bites, mosquitoes are moved from susceptible to an exposed stage of infection with a probability defined by temperature-dependent vector competence as stated in table S2.

In addition, when a household receives a *visitor,* it can spend a random number of hours inside the household between 1 and 3. To account for the difference between the time that inhabitants spend in the house and time *visitors* spend inside, we gave the opportunity to bite a *visitor* to those mosquitoes that were unable to bite the inhabitants on the same day. The probability of unfed mosquitoes to bite the *visitor* is estimated according to probability previously described (*p(b)*).

#### Vector control activities implementation

Four parameters were considered for the vector control strategies in the simulations: 1) the structure type (household, non-household, or both); 2) the presence of containers (small, large, or both); 3) the control intensity (expressed as percentage of eliminated water containers serving as mosquito breeding places: 25%, 50%, 75% and 100%. The latter though it is not intended to be realistic as it is improbable in practice, it is intended to capture a broader range of outcomes for activities); and 4) the day after the beginning of epidemic (only for reactive control: 1, 50, 100 and 250 days, the last only shown in supporting results). In order to implement vector control activities in the simulations, we targeted structures according to environment and presence of containers at the designated day (day 0 for preventive scenario or any of the evaluated under reactive scenario). When the structure met the requirements of the control strategy, the intensity of the control was implemented into a binomial distribution, as follows

$$C_{k}=Bin\left( N_{cont},1-I_{c} \right)$$

where *C_k_* is the number of containers that remains in the structure after the control, *N_cont_* is the total number of containers of the targeted size and *I_c_* is the intensity of the control, expressed as percentage of containers to be eliminated.

#### Model calibration

Dengue in Kenya is not a mandatory reporting disease and its real burden is cloudy by malaria-confounded misdiagnosis [13], so there is not available data to be used as a target for calibration for the entire temporal window modelled. Consequently, we use available seroincidence estimates for 2021 obtained from samples collected on both cities as part of a longitudinal study [14-17]. As part of such study, samples were taken from patients and a second sample was taken six months later. Samples were analyzed with an ELISA following procedures described previously [14, 18, 19]. Seroincidence was estimated as the proportion of positive samples in each month that were negative six months before. To estimate annual rate of infection, we used the equation:

$$rate=-\frac{1}{t}\times\ln\left( 1-p \right)$$

Where the *t* is given in years (since the seroincidence was estimated as seroconversion taking place in six months, the time was set as 0.5 years) and *p* is the proportion of seroconverted individuals. The same formula was applied to results from simulations, where the number of new infections were recorded on the same temporal windows from those estimated from serological data (figure S4).

Ethical approval and oversight for data collection for this study were obtained from the Institutional Review Board of Stanford University (IRB 31488), as well as the Kenya Medical Research Institutes (KEMRI SSC 2611) and Technical University of Mombasa Ethical Review Committee (TUM/ERC EXT/004/2019).

Optimization search was applied on infestation values (final conditions are presented in table S1) starting from those previously reported [1] and on *a* and *b* values of density-dependent function reported in Methods section. Values were sampled as a grid search methodology covering the distribution of biologically plausible ranges. Visual inspection of incidences was used to compare the model versus actual data, Longitudinal and transversal estimates of incidence showing the performance of the model are shown in figure S4.

Calibrated parameters *a* and *b* for the density-dependent function resulted in the following function:

$$f\left( D \right)=\frac{1}{1+e^{0.09D-0.55}}d$$

The study used code to generate the synthetic populations. The code for the model as well as the necessary datasets (which include information about weather, container presence, demographic data, synthetic populations, and distribution parameters) have been deposited at Github site <https://github.com/vhpenagarcia/ABM_dengue>. Files with results of simulations without control and results summary of vector control strategies are also included in the repository.

### Supporting figures

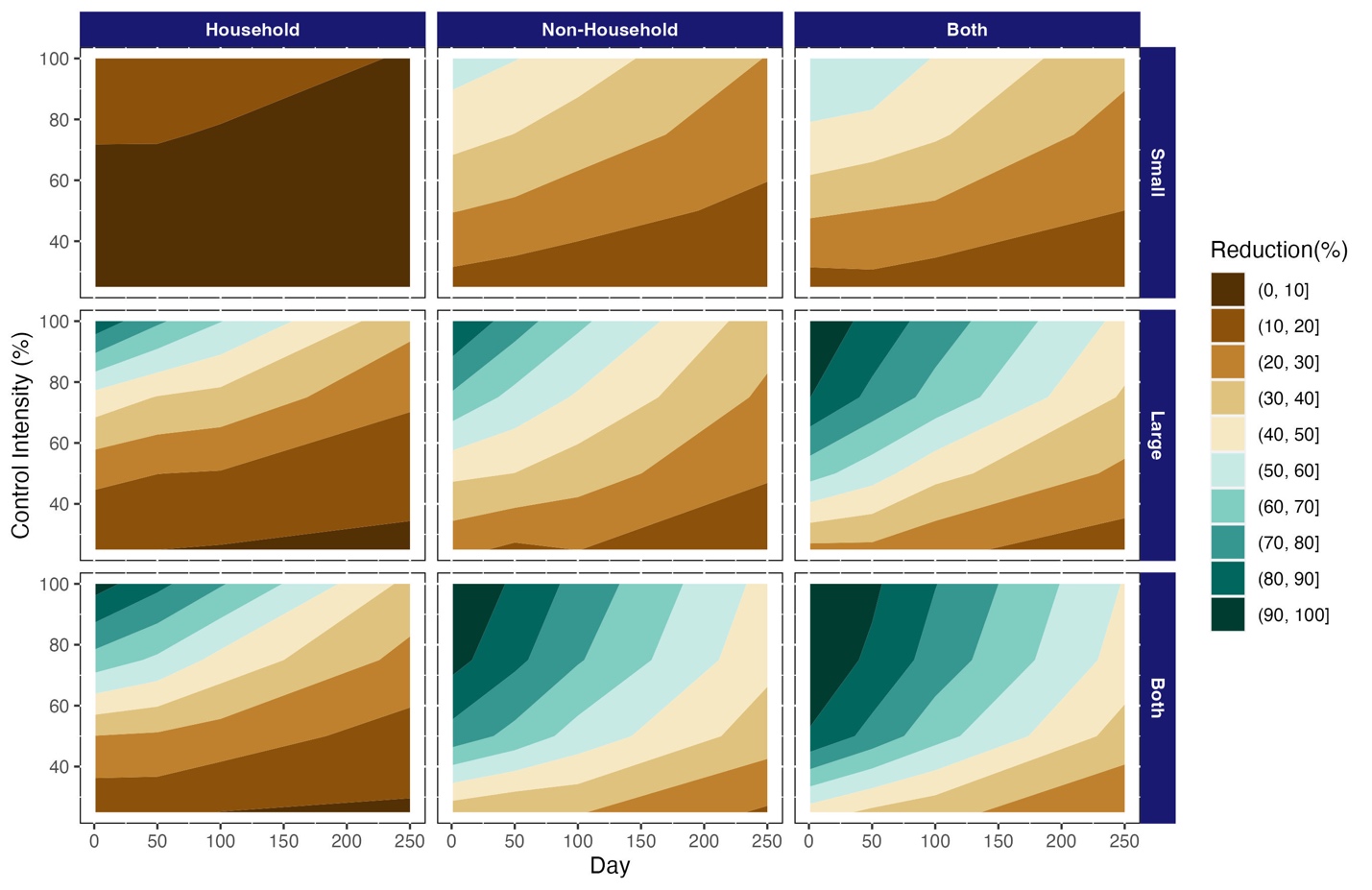

**Figure S2: Reactive control in Ukunda is more effective in NH or all environments combined than HH environments alone, regardless of timing, intensity, container size, and city.** Effectiveness is expressed as the reduction in the percentage of dengue cases compared to the epidemic curve with no vector control interventions. The vector control strategies vary according to control intensity (number of containers eliminated; y-axis), the target environment (Households, Non-Households or both; panel columns); container size (Small, Large or both; panel rows); and the day of implementation after the beginning of the epidemic (x-axis).

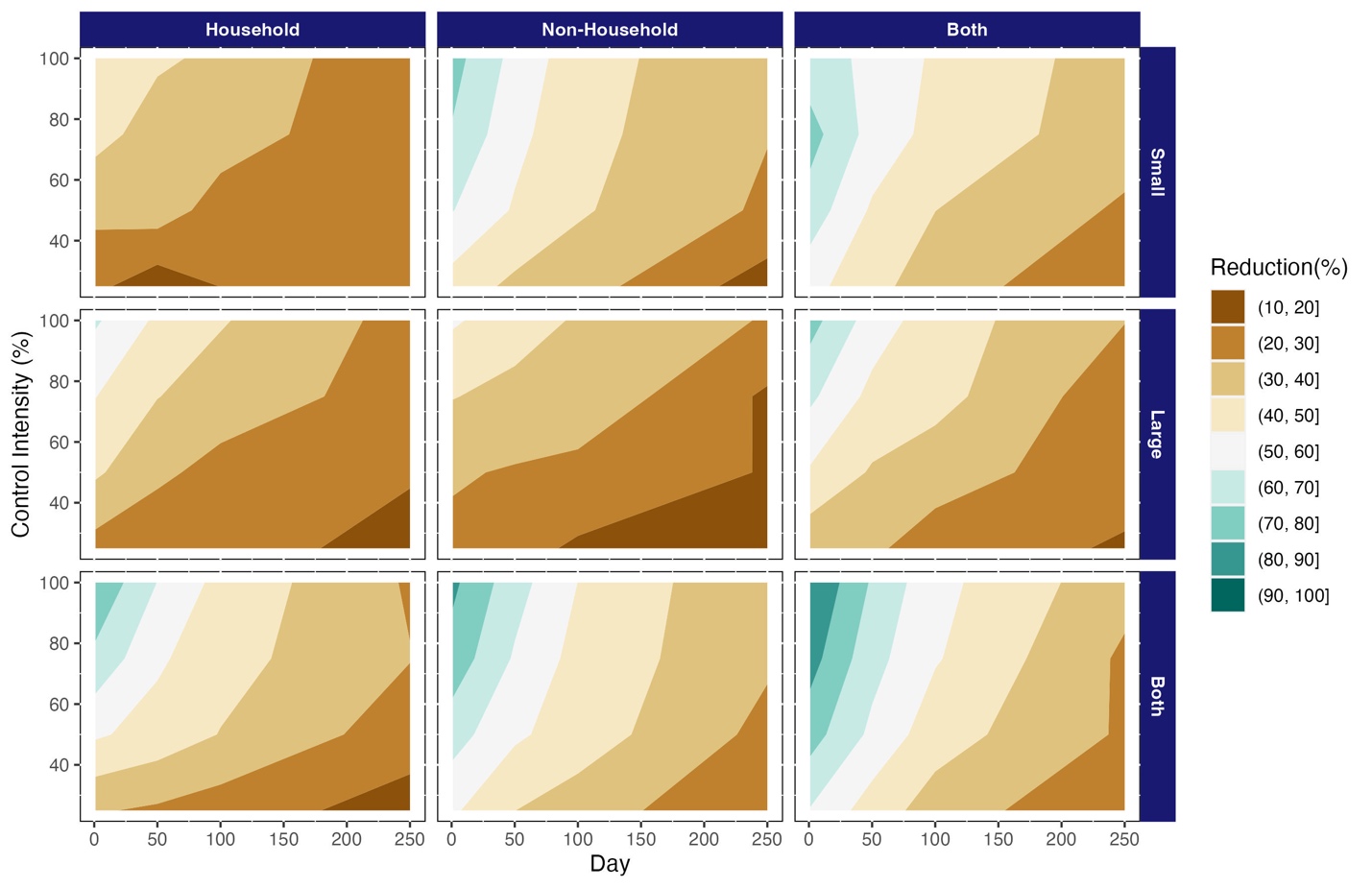

**Figure S3: Reactive control in Kisumu is more effective in NH or all environments combined than HH environments alone, regardless of timing, intensity, container size, and city.** Effectiveness is expressed as the reduction in the percentage of dengue cases compared to the epidemic curve with no vector control interventions. The vector control strategies vary according to control intensity (number of containers eliminated; y-axis), the target environment (Households, Non-Households or both; panel columns); container size (Small, Large or both; panel rows); and the day of implementation after the beginning of the epidemic (x-axis).

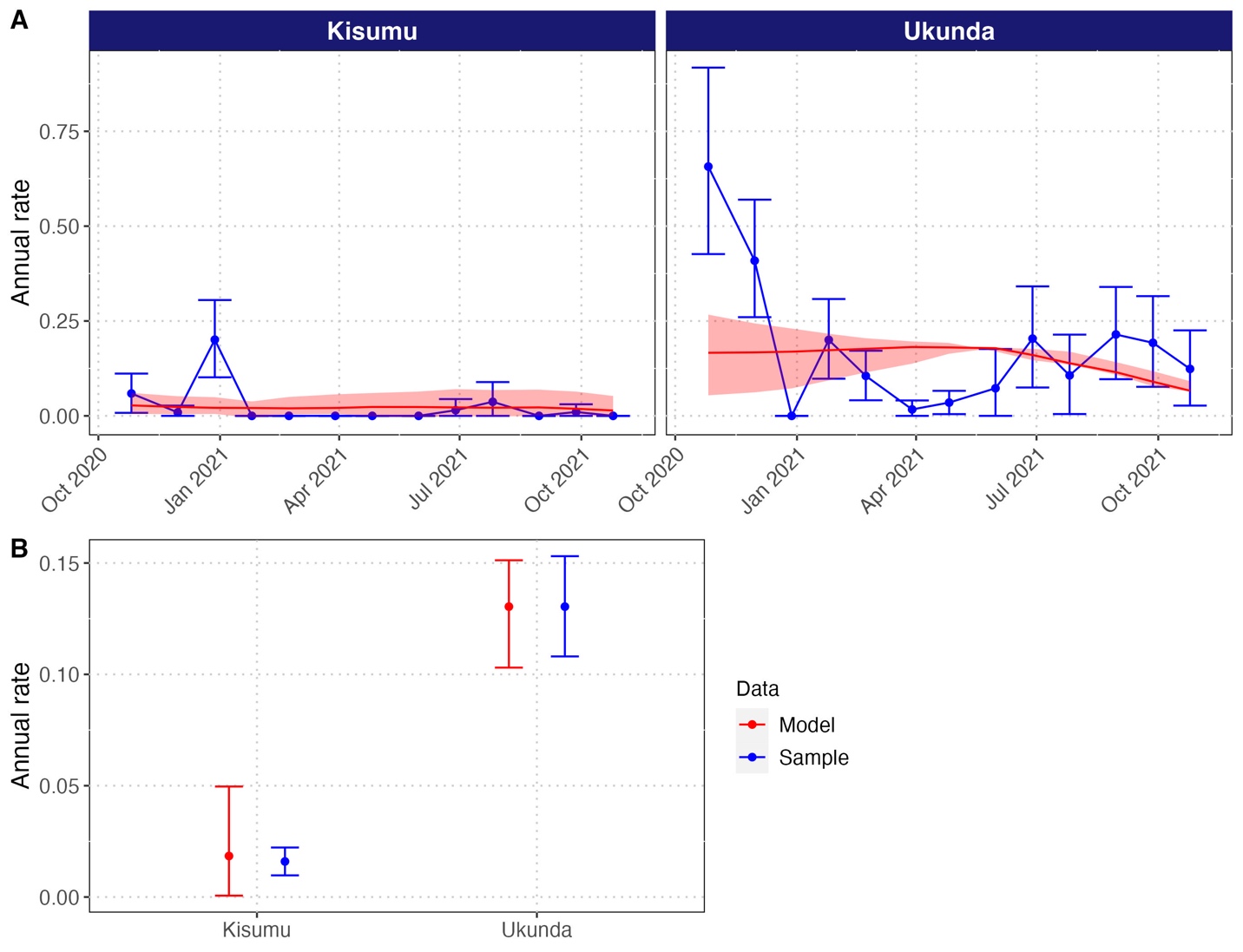

**Figure S4: Results of model calibration process to seroincidence data obtained from samplings done as part of a longitudinal study.** The rates were estimated from proportions calculated from both longitudinal serological samplings and modelled data after calibration. Though data are coming from different sources, the proportions were estimated in a similar way, i.e. proportion of positive individuals from a pool of individuals tested negative six months before. A is depicting the annual rate for both the model and the observed data along sampling time, and B is depicting a single annual rate estimate for the entire sampling.

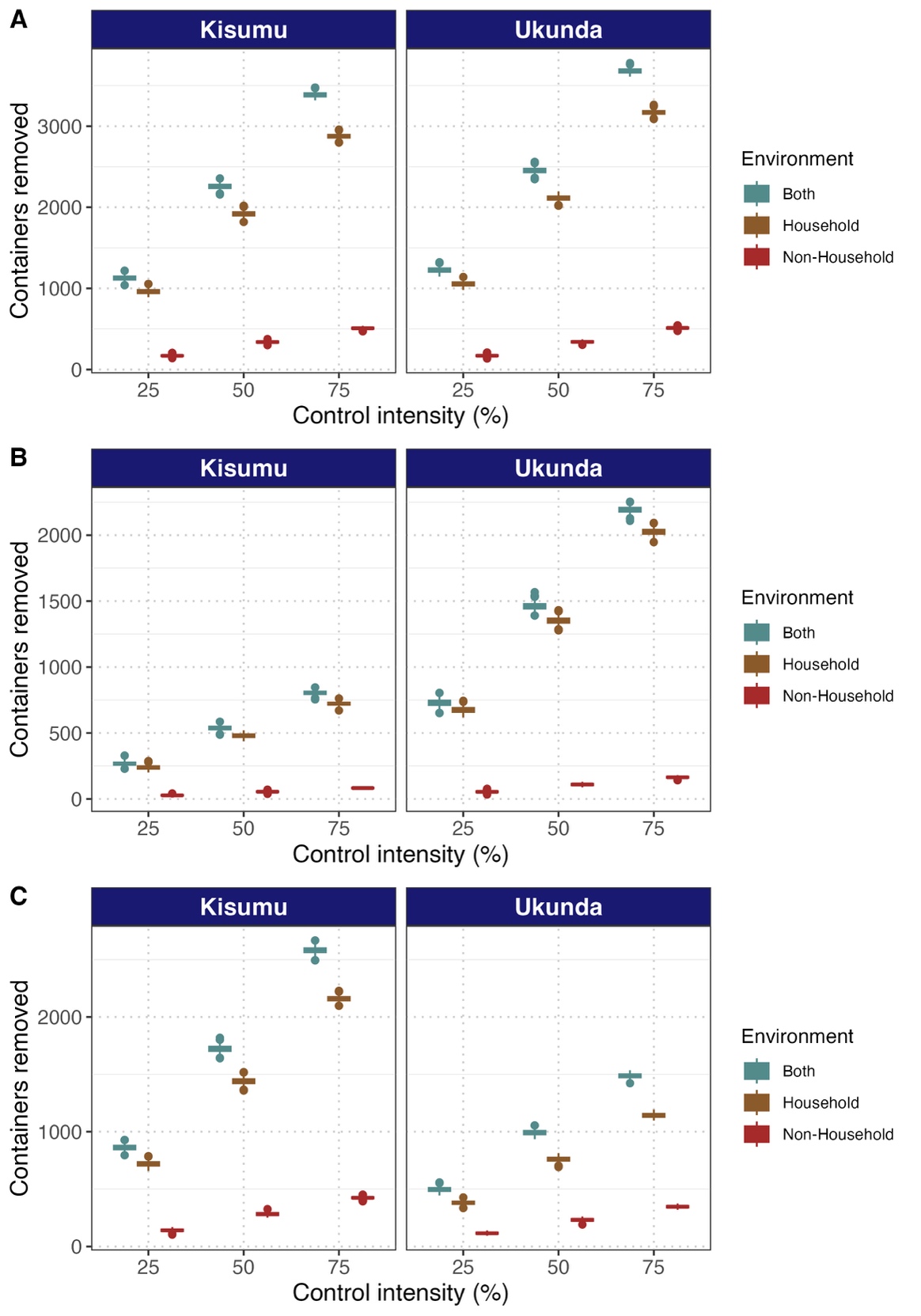

**Figure S5: Number of containers removed in for each assessed strategy for 25, 50, and 75% of control intensity**. Due to its higher number of containers, effort to reduce them in households is higher than when is focusing on either NH or both environments. Plots are showing the number of removed containers when control is applied irrespective of container size (A), on large containers (B), or small containers (C).

### Routine movement questionnaire

The aim of this questionnaire is to collect data on location visited two weeks before case reporting. Given the incubation period of dengue (inoculation → symptoms), the transmission event should have happened at least 7 days before case reporting. At the same time, individuals in infectious status can infect mosquitoes for ~5days. Thus, tracing people movement by identify the most visited locations in the previous two weeks could allow identifying high transmission locations. Although people movement can be acquired by using wearable GPS or mobile phone traffic data, these methods cannot be used to track movement happened in the past weeks. GPS and mobile data are affected by errors due to GPS signal and density of mobile antenna. In order to reduce the error, projects include questionnaires to record the most visited locations of participants and link them with the GPS and mobile data. Our data collection only uses questionnaire because it would not be possible to provide a wearable GPS to all cohort or request mobile data. The questionnaire will be filled by operator trough a ~15 minutes semi-structure interview. During a semi-structured interview, the interviewer will talk with the interviewed person and fill a list of questions. The interviewer will friendly talk with the interviewer and trace back their movement of the last two week using a recalling method based on simple questions about the person common routine. Questions will be focused on common activity that usually people perform in their daily routine. The classical routine activities include working, grocery, school attendance, visiting relatives or friends, attending any cult (such as mass) or entertainment event. For each activity, the interviewer will ask the location in which the activity was performed. For each location, the interviewer will ask landmarks useful for the team to find the location and map it.

#### Questions for the semi-structured interview

The questions should be asked in an informal talk-style interview

##### Adult modules:

1. Do you have a job? (YES: go to question n.2 / NO: go to question n.3)

2. Where do you work? In one particular places or in multiple places? (record selling points/s and landmark to find them, then go to question n.3)

3. Do you sell anything to create income? (YES: go to question n.4 / NO: go to question n.5)

4. Where do you sell your goods? In one particular places or in multiple places? (record selling points/s and landmark to find them, then go to question n.5)

5. Do you usually buy grocery for the house? (YES: go to question n.5 / NO: go to question n.6)

6. Do you buy your grocery at a particular market? (YES: go to question n.7 / NO: go to question n.8)

7. In which market do you usually buy your grocery? In one particular market or in multiple market? (record market/s and landmark to find them, , then go to question n.8)

8. Do you buy your grocery at any shops? (YES: go to question n.9 / NO: go to question n.10)

9. In which shops do you usually buy your grocery? In one particular shop or in multiple shops (record shop/s and landmark to find them, then go to question n.11)

10. Have you met with relatives or friend in a location (houses other type of locations) during the last two weeks? (YES: go to question n.11 / NO: go to question n.13)

12. Could you please tell me where those places are located? (record houses and landmark to find them, then go to question n.13)

13. Have you attended any religion event during the last two week? (YES: go to question n.14 / NO: go to question n.15)

14. Could you please tell me where the religion event was held? (record the religion event location/s and landmark to find them, end of adult module move to child module question n.15)

##### Child module

16. Does the children spend most of his/her time at home? (YES: go to question n.17 / NO: go to question n.17)

17. Does you or someone else in your house (wife, sibling, relative) bring the child with him/her? (YES: go to question n.18 / NO: go to question n. 26)

18. Does this person brings the kid with him/her at work? (YES: go to question n.19 / NO: go to question n.19)

19. Where does this person work? (record work place/s and landmark to find them, then go to question n.20)

20. Does this person bring the child when she/he buys grocery at market? (YES: go to question n.21 / NO: go to question n.22)

21. In which market does the person buys the grocery? In one particular market or in multiple market? (record market/s and landmark to find them, then go to question n.22)

22. Does this person bring the child when she/he buys grocery at any shops? (YES: go to question n.23 / NO: go to question n.24)

23. In which shops does the person buys the grocery? In one particular shop or in multiple shops (record shop/s and landmark to find them, then go to question n.24)

24. Has this person brought the children to a meeting with relatives or friend in a location (houses other type of locations) during the last two weeks? (YES: go to question n.25 / NO: go to question n.26)

25. Could you please tell me where those places are located? (record houses and landmark to find them, then go to question n.26)

26. Has the children attended any religion event during the last two week (alone or brought by someone)? (YES: go to question n.27 / NO: go to question n.28)

27. Could you please tell me where the religion event was held? (record the religion event location/s and landmark to find them, then go to question n.28)

28. Does the child play in other location than his/her house? (YES: go to question n.29 / NO: go to question n.30)

29. Could you please tell us where the child plays? (record the location/s or area/s and landmark to find them, then go to question n.30)

30. Does the child attend school? (YES: go to question n.31 / NO: go to question n.32)

31. Could you please tell us where the school is located? (record the religion event location/s and landmark to find them, then go to question n.32)

[2] Kenya National Bureau of Statistics. 2021 Quarterly Labour Force Report. Quarter 1. (Nairobi.

[3] Kenya Ministry of Education. 2019 Basic Education Statistical Booklet, 2019. In *Basic Education Statistical Booklet* (Nairobi.
